# Socioeconomic position and the influence of food portion size on daily energy intake in adult females: two randomized control trials

**DOI:** 10.1101/2022.12.20.22283727

**Authors:** Tess Langfield, Katie Clarke, Lucile Marty, Andrew Jones, Eric Robinson

## Abstract

**Background:** Large portions of energy-dense foods drive overconsumption and are more readily available in socioeconomically deprived areas. Reducing portion sizes of commercially available foods could be an effective public health strategy to reduce population energy intake, but recent research suggests that the effect portion size has on energy intake may differ based on socioeconomic position (SEP).

**Objective:** We tested whether the effect of reducing food portion sizes on daily energy intake differed based on SEP.

**Methods:** Participants were served either smaller or larger portions of food at lunch and evening meals (N = 50; Study 1) and breakfast, lunch and evening meals (N = 46; Study 2) in the laboratory on two separate days, in repeated-measures designs. The primary outcome was total daily energy intake (kcal). Participant recruitment was stratified by primary indicators of SEP; highest educational qualification (Study 1) and subjective social status (Study 2). Secondary indicators of SEP in both studies included household income, self-reported childhood financial hardship and a measure accounting for total years in education.

**Results:** In both studies, smaller (vs larger) meal portions led to a reduction in daily energy intake (ps < .02). Smaller portions resulted in a reduction of 235kcal per day (95% CI: 134, 336) in Study 1 and 143kcal per day (95% CI: 24, 263) in Study 2. There was no evidence in either study that effects of portion size on energy intake differed by SEP. Results were consistent when examining effects on portion-manipulated meal (as opposed to daily) energy intake.

**Conclusion:** Reducing meal portion sizes could be an effective way to reduce overall daily energy intake and contrary to other suggestions it may be a socioeconomically equitable approach to improving diet. These trials were registered at www.clinicaltrials.gov as NCT05173376 and NCT05399836.

## 1. Introduction

There are well observed socioeconomic inequalities in diet. In developed countries, people from a lower socioeconomic position (SEP) are more likely to consume diets characterised by nutrient-poor, energy-dense foods (1-3), and also have higher rates of obesity (4-6). One potential explanation for socioeconomic disparities in diet and in rates of obesity is the food environment. More deprived areas have a higher density of fast-food outlets in the US (7, 8), and UK (8-10), meaning more widespread availability and ease of access to large portions of energy-dense foods. An additional but related explanation is that the impact the food environment has on diet may differ by SEP, a question we answer in the present research.

A key aspect of the food environment is food portion size. Portion sizes of commercially available foods have increased over time in the US (11) and UK (12). This is concerning given robust evidence that eating from larger compared to smaller portions increases energy intake, a phenomenon known as the ‘portion size effect’ (PSE) (13, 14). A recent meta-analysis revealed that individuals do not fully compensate for changes to portion size over the course of a day (15) and therefore larger portion sizes likely promote increased weight gain (16). Intriguingly, there is some preliminary evidence that the influence of portion size may differ based on SEP, with one study finding that the PSE on hypothetical consumption of unhealthy snacks was larger for participants of lower vs. higher SEP (17). In a different study, lower participant subjective SEP was associated with increased likelihood of eating beyond energy needs after being served a large meal (18), as opposed to eating less energy after the large meal to compensate for increased energy intake. Collectively these studies suggest that the influence of portion size on energy intake may differ by SEP. One interpretation of these findings is that psychological experience of resource scarcity associated with being of lower SEP drives behaviours that promote excess intake in the absence of need, as an energy ‘insurance’ against periods of food scarcity (19-22). However, in another study, participants that were experimentally manipulated to experience a lower socioeconomic position showed a greater sensitivity to the energy content of food (23). This finding was interpreted as being evidence that lower SEP may cause an increased vigilance to the energy content of consumed meals in order to avoid being in energy deprivation. If true, this instead suggests that lower SEP may be associated with an enhanced ability to detect energy intake reductions (e.g., via portion size decreases) which promotes compensation by eating more. Therefore, public health approaches designed to reduce food portion sizes would inadvertently benefit the diet of people from higher SEP more than lower SEP.

As reducing portion size has been identified as a potential target for public health policy (13, 24) and it is critical that approaches do widen existing dietary inequalities (25, 26), further research is needed to understand whether the impact reducing portion sizes has on energy intake differs based on SEP. The present research consists of two laboratory studies examining if the effect of reducing portion size on total daily energy intake is determined by SEP. Consistent with recent research (17, 18), the primary indicators of were highest educational qualification (Study 1) and subjective social status (SSS) (Study 2). Across both studies, a measure accounting for years in higher education and highest educational qualification, income, and self-reported financial hardship in childhood were also examined as additional SEP measures. In both studies, the primary hypothesis was that smaller portions would reduce total daily energy intake and that SEP would moderate the PSE.

## 2. Methods

### 2.1 Participants

The studies were advertised online and in the local community in Liverpool (England). Recruitment materials described the study as investigating ‘mood, diet and sleep’ (cover story). Both studies recruited females aged 18 or over, with a BMI between 18.5-32.5kg/m^2^ (Study 1) and 18.5-39.9kg/m^2^ (Study 2; criteria widened from Study 1 to help boost recruitment). Because of substantial sex differences in energy intake (27, 28), as in (29, 30) we limited studies to females to minimize participant variability in energy intake and therefore increase sensitivity to identify moderating effects of participant level SEP on energy intake. In addition, previous research suggesting SEP moderation of the influence of portion size sampled all (18) or predominantly females (17). Individuals with any dietary restrictions including being vegetarian or dieting (Study 1; though vegetarians were eligible in Study 2 due to study foods), food allergies, self-reported dislike of the test foods, or a history of eating disorders, on mediation affecting appetite or pregnant were ineligible. Participants scoring the middle response (‘5’) on the SSS measure were also excluded in Study 2 as this denotes the midpoint of the SSS measure (i.e., neither high nor low SEP). Participants could not participate in both studies. Recruitment was stratified by the primary SEP indicator for each study (50% lower and higher), as well as by age (50% 18-25 years; 50% 26+ years) in Study 1, and by education (50% <A level or equivalent; 50% >A level or equivalent) in Study 2, to ensure SEP groups differed on the primary SEP indicator but not on other demographics. See Supplementary Materials for full methodological information.

### 2.2 Design

The studies used randomised crossover designs with two study days corresponding to two conditions: smaller vs. larger portion sizes. Study days were on the same day of the week, separated by a washout period of between 1 – 6 weeks. Participants were served all meals in the laboratory and provided additional snacks to take away. Daily energy intake (kcal) was the primary outcome (see **Figure 1** for overview). Participants were randomised to the sequence of conditions using the Microsoft Excel (rand()) function, and were blinded to condition.

**Figure 1.**
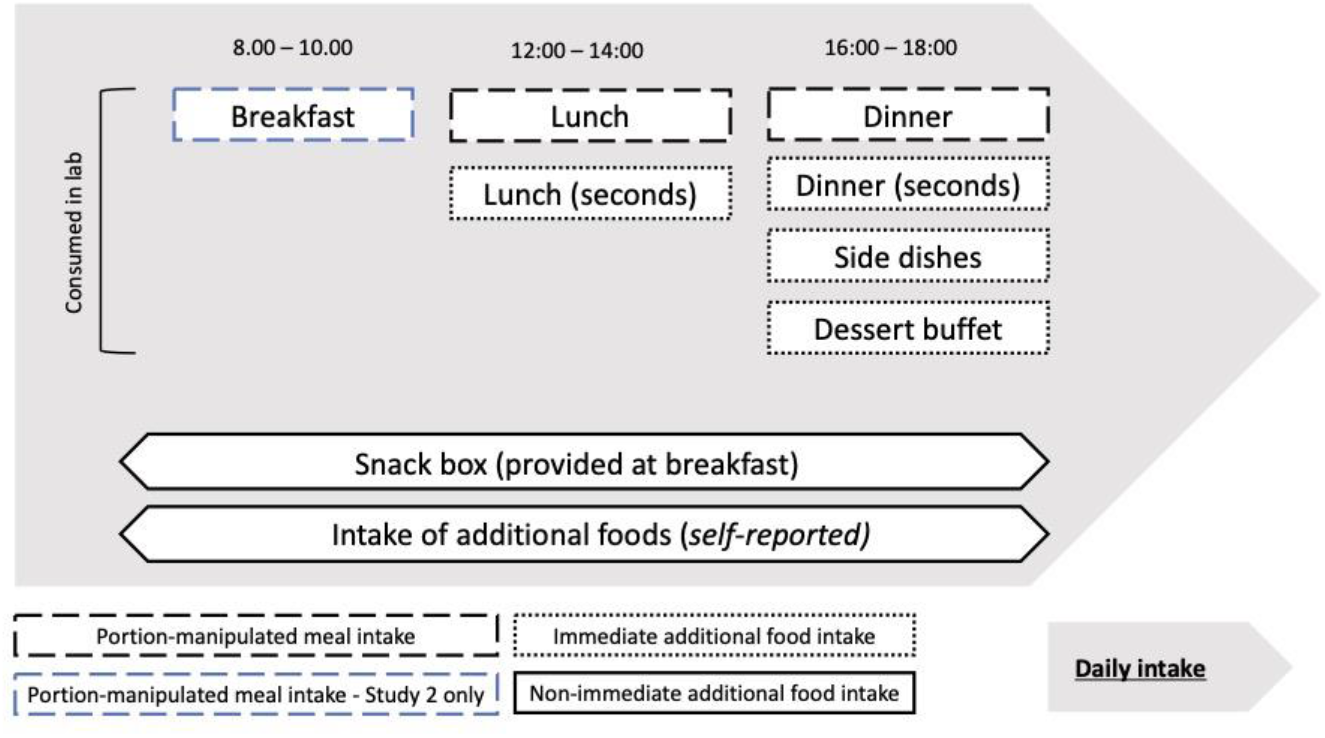
Assessment of daily energy intake (primary outcome) and its components. Diagram adapted from (31).

### 2.3 Study foods

All meal components and snacks were prepared for *ad libitum* consumption. The size of the initial portions served at lunch and dinner (Study 1) and at breakfast, lunch and dinner (Study 2) were manipulated (see **Table 1**). Portion sizes were selected on the basis of previous data (32), so that in both the smaller and larger conditions the amount of food provided appeared a relatively normal meal size, but the larger portions were 50% larger than the smaller portions in g/kcal. As in (31), participants could serve themselves more of the portion manipulated food from large serving bowls, if desired, or request additional servings. See Supplementary Materials for full study menus and macronutrient content of all study food. The dishware used to serve the meals was kept consistent across the two conditions, although in Study 1, the size of breakfast plate and bowl differed based on study day (for more information see Supplementary Materials).

**Table 1.** Portion-manipulated meals.

|  |  | **Portion**<br>**(g)** | **Energy content**<br>**(kcal)** |
| --- | --- | --- | --- |
| **Study 1** | **Lunch** |  |  |
|  | **Cheese and tomato pasta bake** |  |  |
|  | **SMALLER** | 375 | 544 |
|  | **LARGER** | 563 | 816 |
|  | **Dinner** |  |  |
|  | **Beef chilli with rice** |  |  |
|  | **SMALLER** | 291 | 339 |
|  | **LARGER** | 437 | 509 |
| **Study 2** | **Breakfast** |  |  |
|  | **White toast** |  |  |
|  | **SMALLER (2 PIECES)** | 80 | 195 |
|  | **LARGER (3 PIECES)** | 120 | 293 |
|  | **Brown toast** |  |  |
|  | **SMALLER (2 PIECES)** | 80 | 185 |
|  | **LARGER (3 PIECES)** | 120 | 277 |
|  | **Cornflakes** |  |  |
|  | **SMALLER** | 30 | 116 |
|  | **LARGER** | 45 | 174 |
|  | **Yoghurt** |  |  |
|  | **SMALLER** | 100 | 95 |
|  | **LARGER** | 150 | 143 |
|  | **Lunch** |  |  |
|  | **Cheese and tomato pasta bake** |  |  |
|  | **SMALLER** | 350 | 508 |
|  | **LARGER** | 525 | 761 |
|  | **Dinner** |  |  |
|  | **Vegetarian chilli with rice** |  |  |
|  | **SMALLER** | 291 | 296 |
|  | **LARGER** | 437 | 444 |

Notes. Total amount of food available per day including sides and other dishes: 6328kcal (smaller); 6770kcal (larger), in Study 1 and 5040kcal (smaller); 5643kcal (larger), in Study 2.

### 2.4 Participant measures

#### 2.4.1 Highest education level

In Study 1, highest educational qualification was the primary SEP indicator. As in (17) participants with A-levels or less were categorised as lower SEP, and those with higher education (e.g. degree level) as higher SEP.

#### 2.4.2 Subjective socioeconomic status (SSS)

In Study 2, SSS was the primary SEP indicator. SSS was measured using the MacArthur Scale, which probes an individual’s perceived position in society relative to others in terms of money, education, and jobs. Participants were asked: “*Think of a ladder as representing where people stand in society. At the top of the ladder, are the people who are best off — those who have the most money, most education, and the best jobs. At the bottom, are the people who are worst off—who have the least money, least education and the worst jobs or no job. The higher up you are on this ladder, the closer you are to people at the very top and the lower you are, the closer you are to the bottom. Please select where best represents where you think you stand on the ladder*.” The ladder scored from 1 – 10, with higher scores indicating higher perceived social status (33). Participants scoring 1 – 4 were categorised as lower SEP and those scoring 6 – 10 as higher SEP, with individuals scoring 5 excluded to ensure the two groups clearly differed in SSS.

#### 2.4.3 Other socioeconomic indicators

Additional measures of SEP were collected for use in sensitivity analyses. To account for both education qualifications achieved and time in education, ‘Level of education’ was calculated from the self-reported number of years spent in higher education and highest educational qualification (coded 1 – 9), which were each z-scored and averaged to produce a composite score. Equivalised household income was derived from self-reported household income adjusted for household composition (34). Self-reported financial hardship in childhood was measured using a 3-item resource availability questionnaire (e.g. *“*My family had enough money for things growing up*”*), coded from 1 [strongly disagree] – 7 [strongly agree], with scores averaged, and lower scores indicating greater financial hardship (lower resource availability) (35).

#### 2.4.4 Individual difference measures

To potentially explain any SEP differences in responsiveness to the portion-size effect we collected a range of trait measures. We measured self-reported impulsivity, inhibition (Stroop task), health and weight control food choice motives, satiety responsiveness, plate-clearing tendencies, perceived food insecurity, compensatory health beliefs, and perceived ‘normal’ portion sizes. See Supplementary Materials for full information.

### 2.5 Study outcome measures

#### 2.5.1 Daily energy intake

Energy intake (kcal) was assessed by weighing study food before and after consumption using digital scales ([Sartorius], measured to the nearest 0.1 g). Consumed weight (g) was multiplied by energy density for each food item (kcal/g) derived from the food packaging. Any energy intake (kcal) from self-reported non-study food energy intake (kcal) was estimated using intake24, a validated online dietary assessment tool (36, 37).

#### 2.5.2 Energy intake from foods with portion manipulation

Energy (kcal) from portion-manipulated food items only (i.e., initial portion-manipulated servings).

#### 2.5.3 Energy intake from non-manipulated foods

Energy (kcal) from food and drinks which were not portion-manipulated (i.e., additional servings/seconds, sides, desserts, snacks, and self-reported additional food/drink).

#### 2.5.4 Physical activity

To assess compliance with study instructions (to avoid vigorous physical exercise and to keep physical activity roughly consistent on both study days), participants wore activity monitors on each study day (Fitbit Zip). Data on moderate-vigorous activity (operationalised as total minutes of activity with a metabolic equivalent of ≥3) was collected. Fitbit device active minute estimates have been validated against gold standard research-grade physical activity monitoring devices (38).

#### 2.5.5 Hunger and fullness

Pre- and post-meal hunger and fullness were assessed as part of a battery of filler ‘mood’ measures using visual analog scales ranging from 0 (‘not at all’) to 100 (‘extremely’), with 12 measures in total: [breakfast, lunch, dinner] x [pre-meal, post-meal] x [smaller portion study day, larger portion study day]. Hunger and fullness ratings were plotted separately for each condition and the area under the curve (AUC) was calculated using the trapezoid function (39).

#### 2.5.6 Liking, familiarity, normality

For all study foods, participants rated liking (1 [not at all] -7 [very much]), and familiarity (’I would normally eat this type of food’; 1 [strongly disagree] – 7 [strongly agree]). Participants were also shown images of each portion for the portion-manipulated foods and asked how ‘normal’ it was (1 [not normal, it is far too small] – 7 [not normal, it is far too big]).

#### 2.5.7 Awareness of study aim and portion manipulation

Participants described what they believed the aim of the study was, in an open-ended text response, with two researchers independently assessing awareness (i.e., the impact of portion size on energy intake). Participants were then asked whether they noticed the difference in portion size between study days, and which portion they had on each study day. Participants were coded as ‘aware’ of the portion size manipulation if they answered ‘yes’ to the first question, and correctly identified the order in which they received smaller vs. larger portions.

### 2.6 Procedure

Participants completed a screening session in which eligibility was confirmed, weight and height were measured, and two individual difference measures were completed (Stroop task and perceived ‘normal’ portion size task). Prior to study days participants were asked to avoid eating or drinking anything other than water before attending for breakfast, and to not eat anything other than the provided study food (teas, coffees, water, soft drinks and up to 2 alcoholic beverages could be consumed as normal, but we asked that they recorded them as additional food/drink). To bolster the cover story, participants completed filler mood questions before and after eating each meal (with embedded hunger and fullness ratings), a filler word-categorisation task relating to ‘mood’ words during lunch, and a questionnaire about their sleep during breakfast. Meals were served onto a dining table, with non-manipulated food (e.g., additional serving of the portion size manipulated food, sides, dessert) placed on an adjacent serving table. All meals were served with 500ml chilled water, and a choice of tea or coffee with breakfast. When being served each meal, participants were told they could eat as much or as little as they would like, and that they were not time-limited. At the end of each breakfast session, participants were provided with a snack box to take away and consume food from when required, a Fitbit activity monitor, and food diary to record any drinks or additional food consumed throughout the day. After attending both study days participants attended a final session during which they completed a battery of questionnaires (i.e., individual difference measures) and a study experience questionnaire (i.e., probing awareness of the aim of the study), before being debriefed. Participants were reimbursed for their time and any travel expenses. See Supplementary Materials for full methodological information.

Study 1 was conducted between October 2021 and April 2022 and Study 2 was conducted between June and October 2022. Both studies were conducted in line with institutional ethical guidelines and received ethical approval (reference: 6154). Study protocols were pre-registered on the Open Science Framework (Study 1: osf.io/gkrp7; Study 2: osf.io/hx75k) and on ClinicalTrials.gov (Study 1: NCT05173376; Study 2: NCT05399836).

### 2.7 Analysis plan

All analyses were pre-registered unless stated. Analyses were conducted using SPSS version 26 (frequentist analyses) and JASP version 0.16.4 (Bayesian analyses). Main effects and interactions were assessed against an alpha of p < .05 for primary and p < .01 for secondary analyses to account for multiple comparisons. Bayes Factors were computed using default priors; r scale fixed effects = 0.5, r scale random effects = 1, and r scale covariates = 0.353. We report BFincl and use conventional cut offs as evidence for the alternative hypothesis (1 – 3: anecdotal; 3 – 10: moderate; 10 – 30: strong; 30 – 100 very strong; > 100 extreme; with inverse values indicative of the same degree of evidence for the null) (40).

#### 2.7.1 Sample size

Power calculations were conducted using G*Power 3.1. For Study 1, we calculated that a minimum sample of 46 participants was required to detect a small-to-medium main effect of portion size or interaction with SEP on daily energy intake (f=0.175, p < .05, 85% power, within-subject correlation of 0.7 estimated from (31)). We powered Study 2 to detect the same sized effects and interactions on daily energy intake (f=0.175, p < .05, 85% power, within-subject correlation of 0.75 (conservatively used given Study 1 r = 0.84]), resulting in a minimum N=40. As specified in the protocol, we planned to recruit slightly above minimum sample sizes (10-15%) to account for any missing data and maximise statistical power. Recruited n = 54 (Study 1) and n = 47 (Study 2).

#### 2.7.2 Primary analyses

The primary analyses were mixed ANOVA testing the within-subjects effects of portion size (smaller, larger), between-subjects effects of SEP (lower vs higher), and the interaction (portion size*SEP) on daily energy intake. Bayes Factors were computed for all main effects and interactions.

#### 2.7.3 Sensitivity analyses

We tested the sensitivity of the primary analyses by repeating the analyses after: i) excluding participants guessing the aims of the study, ii) excluding outliers (identified as those with a value > 3SD from condition mean) and influential cases (identified as those with a Cook’s distance > 1, indicating a multivariate outlier (41)), iii) adjusting for portion size order (smaller first, larger first), and iv) substituting the primary socioeconomic indicator with alternative measures: level of education, equivalised household income, SSS (continuous measure), self-reported financial hardship in childhood (continuous measure, pre-registered Study 2 only), and highest educational qualification (Study 2 only).

#### 2.7.4 Secondary analyses

We ran four mixed ANOVAs testing the within-subjects effects of portion size (smaller, larger), between-subjects effects of SEP (lower vs higher), and the interaction (portion size*SEP) on i) energy intake from foods with portion manipulation, and ii) energy intake from non-manipulated foods (analyses pre-registered for Study 2 only), iii) AUC for hunger, and iv) AUC for fullness (to assess whether changes to portion size were associated with changes in hunger and fullness). We also examined if minutes of moderate-vigorous physical activity differed between SEP or portion size conditions (see Supplementary Materials). Finally, liking, familiarity and normality ratings are reported for each portion-manipulated dish.

## 3. Results

### 3.1 Sample characteristics

For Study 1, the final analysis sample was N = 50 and for Study 2 the final sample was N = 46; see **Figure 2** for participant flow diagram and exclusions. There was no evidence in Study 1 or 2 that SEP groups differed in age (ps > .159) or BMI (ps > .125). In Study 1, the higher SEP group had more current students (university) than the lower SEP. See **Table 2** for summary participant characteristics.

**Table 2.** Summary participant characteristics split by SEP group and overall

|  | Study 1 |  |  | Study 2 |  |  |
| --- | --- | --- | --- | --- | --- | --- |
|  | Lower SEP<br>(n = 25) | Higher SEP<br>(n = 25) | Overall<br>(N = 50) | Lower SEP<br>(n = 23) | Higher SEP<br>(n = 23) | Overall<br>(N = 46) |
| **Age** | 46.36 (18.35) | 38.20 (19.88) | 42.28 (19.37) | 50.00 (17.74) | 53.13 (13.28) | 51.57 (15.58) |
| **Ethnic group** |  |  |  |  |  |  |
| White | 24 (96%) | 18 (72%) | 42 (84%) | 21 (91.3%) | 22 (95.7%) | 43 (93.5%) |
| Mixed or multiple | - | 1 (4%) | 1 (2%) | - | 1 (4.3%) | 1 (2.2%) |
| Asian or Asian British | - | 6 (24%) | 6 (12%) | 1 (4.3%) | - | 1 (2.2%) |
| Black, African, Caribbean, or Black British | 1 (4%) | - | 1 (2%) | 1 (4.3%) | - | 1 (2.2%) |
| Other ethnic group | - | - | - | - | - | - |
| **Student or employment status** |  |  |  |  |  |  |
| Current student | 3 (12%) | 12 (48%) | 15 (30%) | 2 (8.7%) | 1 (4.3%) | 3 (6.5%) |
| Full or part time | 10 (40%) | 6 (24%) | 16 (32%) | 9 (39.1%) | 12 (52.2%) | 21 (45.7%) |
| Looking after home/family | - | 1 (4%) | 1 (2%) | 3 (13%) | 2 (8.7%) | 5 (10.9%) |
| Retired | 9 (36%) | 6 (24%) | 15 (30%) | 5 (21.7%) | 8 (34.8%) | 13 (28.3%) |
| Unemployed/other | 3 (12%) | - | 3 (6%) | 2 (8.7%) | - | 2 (4.3%) |
| Temporary or permanently sick or disabled | - | - | - | 2 (8.7%) | - | 2 (4.3%) |
| **Highest educational qualification achieved or currently working towards** |  |  |  |  |  |  |
| No formal qualifications | 2 (8%) | - | 2 (4%) | 1 (4.3%) | 1 (4.3%) | 2 (4.3%) |
| 1-3 GCSEs or equivalent – *US equivalent: High School Diploma/GED Certificate* | 2 (8%) | - | 2 (4%) | 2 (8.6%) | 1 (4.3%) | 3 (6.5%) |
| 4+ GCSEs or equivalent – *US equivalent: High School Diploma/GED Certificate* | 9 (36%) | - | 9 (18%) | 4 (17.5%) | 5 (21.7%) | 9 (19.6%) |
| A level or equivalent – *US equivalent: Advanced Placement* | 12 (48%) | - | 12 (24%) | 4 (17.4%) | 4 (17.4%) | 8 (17.4%) |
| Certificate of higher education (CertHE) or equivalent – *US equivalent: Associate degree* | - | 2 (8%) | 2 (4%) | 1 (4.3%) | - | 1 (2.2%) |
| Diploma of higher education (DipHE) or equivalent | - | 4 (16%) | 4 (8%) | - | 2 (8.7%) | 2 (4.3%) |
| Bachelor or equivalent | - | 12 (48%) | 12 (24%) | 10 (43.5%) | 6 (26.1%) | 16 (34.8%) |
| Master's degree or equivalent | - | 6 (24%) | 6 (12%) | 1 (4.3%) | 4 (17.4%) | 5 (10.9%) |
| Doctorate or equivalent | - | 1 (4%) | 1 (2%) | - | - | - |
| **Years in higher education** | 1.20 (1.14) | 4.80 (3.03) | 3.00 (2.96) | 2.83 (2.49) | 3.39 (2.45) | 3.11 (2.46) |
| **Level of education (composite z score)** | -.40 (.89) | .41 (.58) | .008 (.85) | -0.102 (0.973) | 0.052 (0.998) | -0.025 (0.98) |
| **Equivalised household income (£)<sup>a</sup>** | 14332.22<br>(8216.25) | 20166.88<br>(15878.25) | 17314.38<br>(12922.01) | 14129.25<br>(7896.04) | 25321.66<br>(14801.25) | 19868.95<br>(13084.06) |
| **Subjective socioeconomic status (1 – 10)** | 5.32 (1.65) | 5.68 (1.11) | 5.50 (1.40) | 3.43 (0.662) | 6.65 (0.775) | 5.04 (1.78) |
| **BMI (kg/m<sup>2</sup>)** | 26.50 (3.66) | 25.18 (2.07) | 25.84 (3.02) | 27.65 (4.83) | 28.30 (5.69) | 27.98 (5.23) |
| **Weight status** |  |  |  |  |  |  |
| Normal weight | 10 (40%) | 11 (44%) | 21 (42%) | 9 (39.1%) | 7 (30.4%) | 16 (34.8%) |
| Overweight | 11 (44%) | 13 (52%) | 24 (48%) | 7 (30.4%) | 10 (43.5%) | 17 (37.0%) |
| Class I obesity | 4 (16%) | 1 (4%) | 5 (10%) | 4 (17.4%) | 2 (8.7%) | 6 (13.0%) |
| Class II obesity | - | - | - | 3 (13.1%) | 4 (17.4%) | 7 (15.2%) |

Notes. Values are M(SD), or counts (%). SEP: socioeconomic position.<sup>a</sup> Study 1 data from sample of n = 45 and Study 2 data from sample of n = 39 (individuals with implausible or missing data excluded; n = 5 in Study 1 and n = 7 in Study 2). <sup>b</sup> higher scores indicate higher resource availability. Further educational equivalents are reported in the Supplementary Materials.

**Figure 2.**
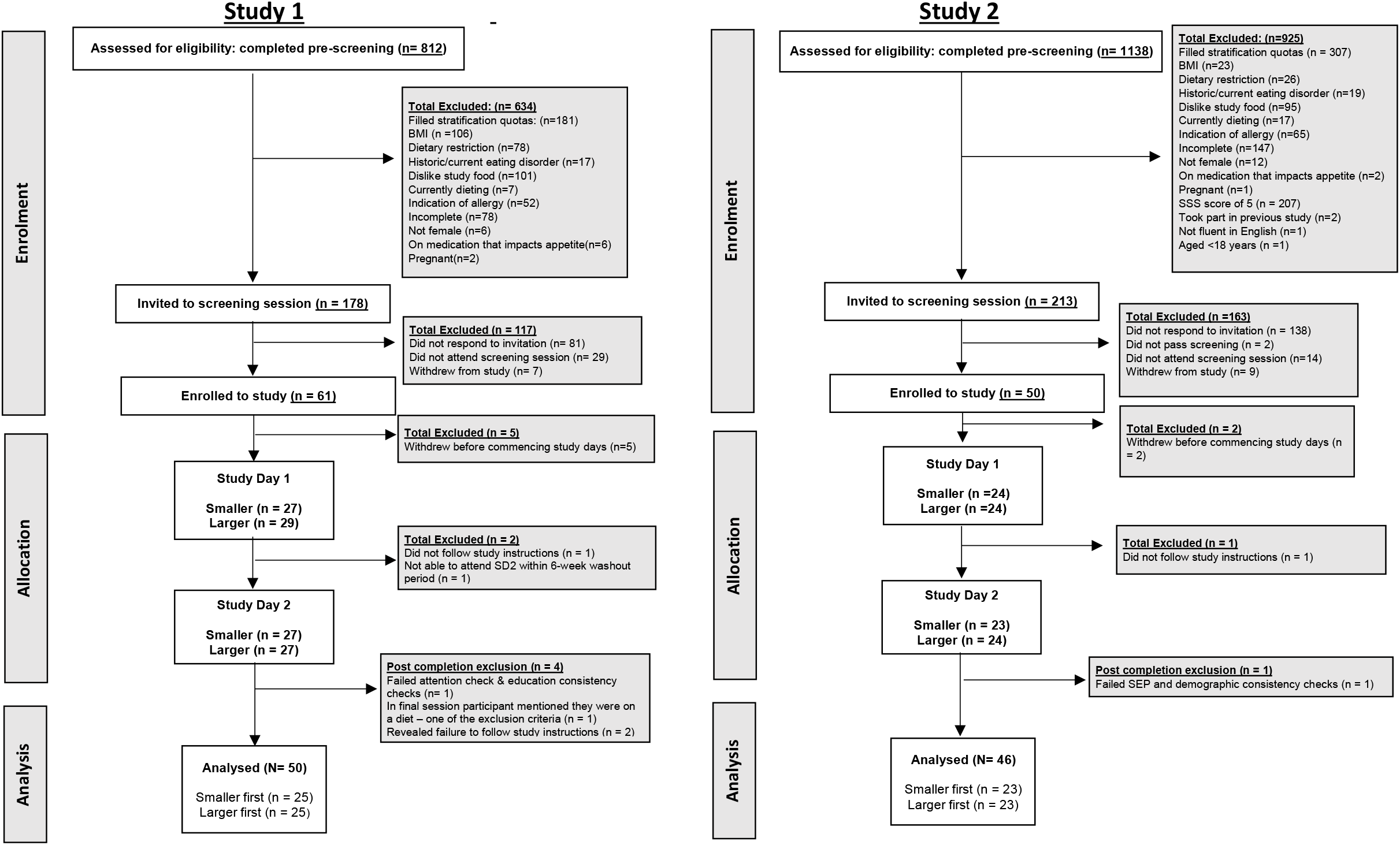
CONSORT flowchart for participant enrolment, allocation, and analysis for Study 1 (left panel) and Study 2 (right panel). Attention checks were included in online questionnaires (e.g., “When did you last visit the Moon”). Consistency checks were also included in online questionnaires (e.g., verifying highest educational qualification).

### 3.2 Effect of portion size on daily energy intake

#### 3.2.1 Study 1

In Study 1, when smaller portions were served, daily energy intake was 235kcal lower (95% CI: 134, 336) vs. larger portions. Higher SEP individuals ate 426kcal more than lower SEP individuals (95% CI: 114, 739), but there was no evidence that SEP moderated the portion size effect, see **Figure 3**. The Bayes factor for the main effect of portion size was BF10 > 100, indicative of extreme evidence for the alternative hypothesis (i.e., smaller meal portions decrease daily energy intake). The Bayes factor for the main effect of SEP was BF10 = 4.7 indicative of moderate support for the alternative hypothesis (i.e., daily energy intake higher among higher SEP vs lower SEP). The Bayes factor for the portion size*SEP interaction was BF10 = 1.01, indicative of no clear evidence. When statistically adjusting for student status the main effect of SEP was no longer significant suggesting the main effect of SEP was being driven by student status (there were more current students in the higher SEP group; see Supplementary Materials).

**Figure 3.**
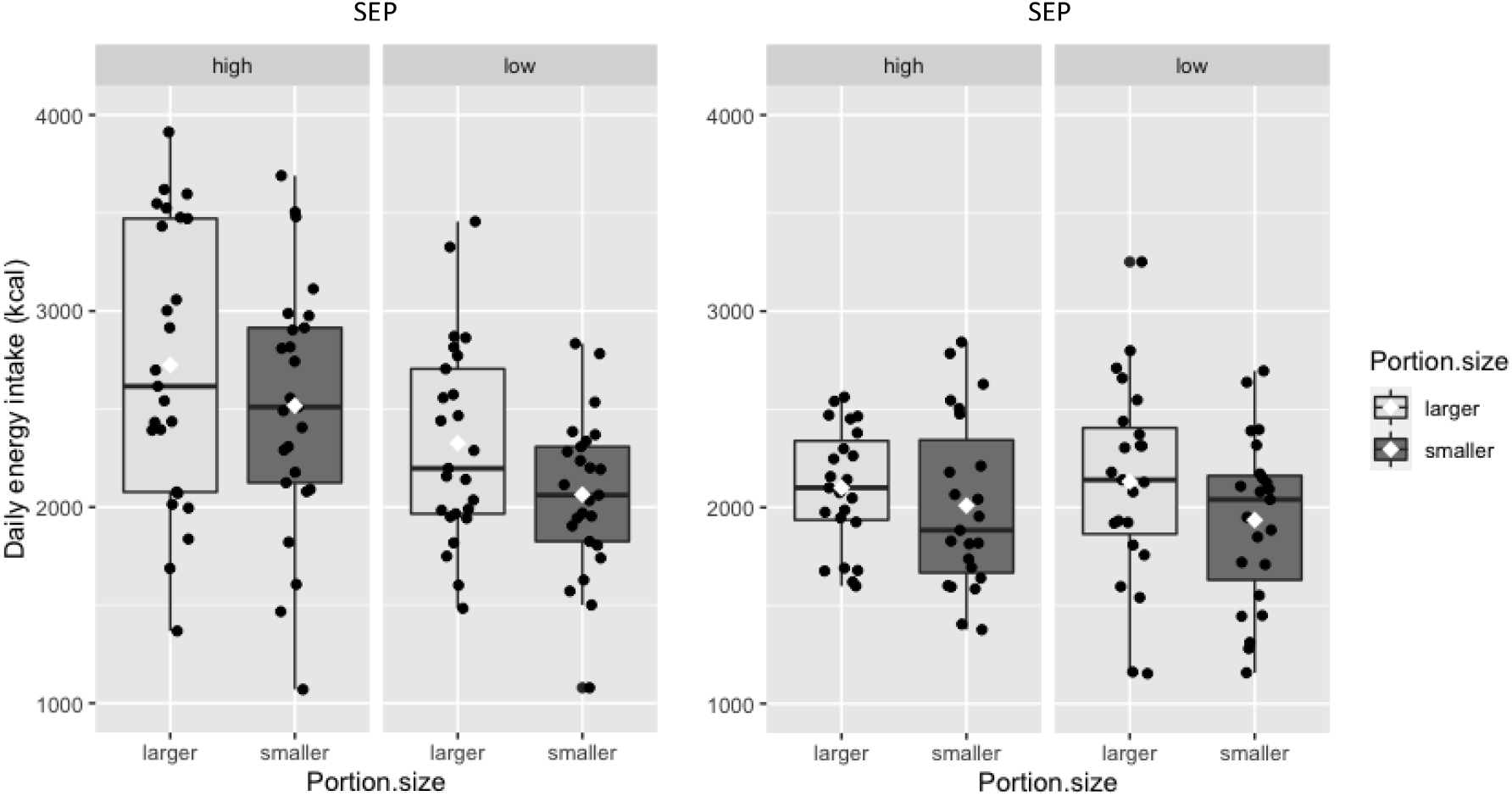
Daily energy intake by socioeconomic position (SEP) and portion size condition for Study 1 (left) and Study 2 (right). Boxplots with median (IQR), with means denoted by white diamonds.

#### 3.2.2 Study 2

The same pattern of results was observed in Study 2; when smaller portions were served, daily energy intake was 143kcal lower (95% CI: 24, 263) vs. larger portions. There was no evidence that SEP groups differed on energy intake, and no evidence that SEP moderated the portion size effect, see Figure 3. The Bayes factor for the main effect of portion size was BF10 =1.854, indicative of anecdotal evidence for the alternative hypothesis (i.e. smaller portions decrease energy intake). The Bayes factor for the main effect of SEP was BF10 = 0.308 indicative of moderate support for the null hypothesis (i.e., no difference in energy intake between higher SEP vs lower SEP). Critically, the Bayes factor for the portion size*SEP interaction was BF10 = 0.305, indicative of moderate support for the null hypothesis.

For energy intake data and full statistical models see **Tables 3 and 4** respectively. In all sensitivity analyses the pattern of findings did not differ, whereby the interaction between portion size and SEP on daily energy intake remained non-significant. See Supplementary Materials for full results and Supplementary Table S6 for full ANOVA results for SEP sensitivity analyses.

**Table 3.** Study outcome measures by portion size condition and SEP group (M, SD)

|  | Study 1 |  |  |  | Study 2 |  |  |  |
| --- | --- | --- | --- | --- | --- | --- | --- | --- |
|  | Higher SEP |  | Lower SEP |  | Higher SEP |  | Lower SEP |  |
|  | Smaller | Larger | Smaller | Larger | Smaller | Larger | Smaller | Larger |
| **Total daily energy intake (kcal)** | 2517.44 (641.12) | 2724.83 (711.90) | 2063.59 (397.32) | 2326.05 (509.51) | 2009.261 (438.11) | 2099.951 (2099.95) | 1935.548 (427.29) | 2131.757 (506.51) |
| **Energy intake from portion-manipulated food (kcal)** | 794.61 (98.81) | 1074.74 (225.58) | 813.08 (84.30) | 1030.80 (187.18) | 1013.52 (199.64) | 1179.53 (262.32) | 993.91 (176.28) | 1230.96 (271.99) |
| **Energy intake from non-manipulated foods (kcal)** | 1722.8 (595.17) | 1650.10 (631.66) | 1250.5 (361.19) | 1295.24 (403.65) | 995.74 (315.87) | 920.42 (246.31) | 941.63 (375.63) | 900.79 (390.97) |
| **Hunger<sup>a</sup>** | 150.80 (62.14) | 149.74 (61.89) | 133.38 (47.81) | 135.68 (53.21) | 174.7 (54.6) | 177.2 (67.8) | 171.3 (71.9) | 159.5 (67.0) |
| **Fullness<sup>a</sup>** | 273.10 (55.98) | 287.40 (58.22) | 270.66 (55.43) | 277.86 (49.55) | 267.0 (49.8) | 274.7 (46.6) | 267.5 (67.9) | 276.8 (59.6) |

Notes. Values are M(SD). SEP: socioeconomic position. Foods with portion manipulation for Study 1: lunch (initial portion), dinner (initial portion), for Study 2: breakfast (initial portions), lunch (initial portion), dinner (initial portion). <sup>a</sup> Area under the curve of meal ratings taken before and after each meal, across entire day.

**Table 4.**
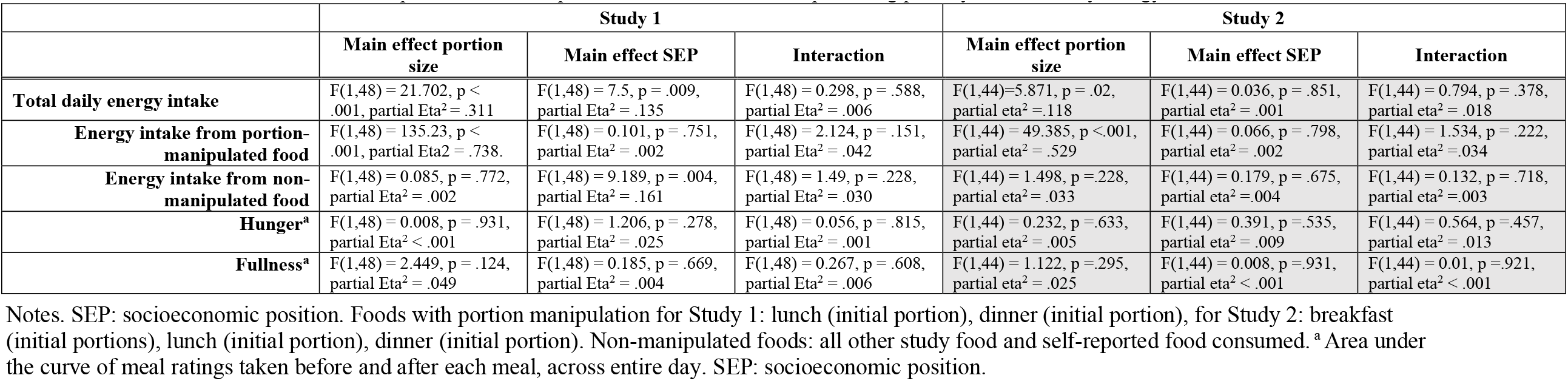
Mixed ANOVA results: portion size, SEP, portion size*SEP interaction, predicting primary and secondary energy intake outcomes

|  | Study 1 |  |  | Study 2 |  |  |
| --- | --- | --- | --- | --- | --- | --- |
|  | Main effect portion size | Main effect SEP | Interaction | Main effect portion size | Main effect SEP | Interaction |
| **Total daily energy intake** | F(1,48) = 21.702, p < .001, partial Eta<sup>2</sup> = .311 | F(1,48) = 7.5, p = .009, partial Eta<sup>2</sup> = .135 | F(1,48) = 0.298, p = .588, partial Eta<sup>2</sup> = .006 | F(1,44)=5.871, p = .02, partial eta<sup>2</sup> =.118 | F(1,44) = 0.036, p = .851, partial eta<sup>2</sup> = .001 | F(1,44) = 0.794, p = .378, partial eta<sup>2</sup> = .018 |
| **Energy intake from portion-manipulated food** | F(1,48) = 135.23, p < .001, partial Eta<sup>2</sup> = .738. | F(1,48) = 0.101, p = .751, partial Eta<sup>2</sup> = .002 | F(1,48) = 2.124, p = .151, partial Eta<sup>2</sup> = .042 | F(1,44) = 49.385, p <.001, partial eta<sup>2</sup> = .529 | F(1,44) = 0.066, p = .798, partial eta<sup>2</sup> = .002 | F(1,44) = 1.534, p = .222, partial eta<sup>2</sup> =.034 |
| **Energy intake from non-manipulated food** | F(1,48) = 0.085, p = .772, partial Eta<sup>2</sup> = .002 | F(1,48) = 9.189, p = .004, partial Eta<sup>2</sup> = .161 | F(1,48) = 1.49, p = .228, partial Eta<sup>2</sup> = .030 | F(1,44) = 1.498, p =.228, partial eta<sup>2</sup> = .033 | F(1,44) = 0.179, p = .675, partial eta<sup>2</sup> =.004 | F(1,44) = 0.132, p = .718, partial eta<sup>2</sup> =.003 |
| **Hunger<sup>a</sup>** | F(1,48) = 0.008, p = .931, partial Eta<sup>2</sup> < .001 | F(1,48) = 1.206, p = .278, partial Eta<sup>2</sup> = .025 | F(1,48) = 0.056, p = .815, partial Eta<sup>2</sup> = .001 | F(1,44) = 0.232, p =.633, partial eta<sup>2</sup> = .005 | F(1,44) = 0.391, p =.535, partial eta<sup>2</sup> = .009 | F(1,44) = 0.564, p =.457, partial eta<sup>2</sup> = .013 |
| **Fullness<sup>a</sup>** | F(1,48) = 2.449, p = .124, partial Eta<sup>2</sup> = .049 | F(1,48) = 0.185, p = .669, partial Eta<sup>2</sup> = .004 | F(1,48) = 0.267, p = .608, partial Eta<sup>2</sup> = .006 | F(1,44) = 1.122, p =.295, partial eta<sup>2</sup> = .025 | F(1,44) = 0.008, p =.931, partial eta<sup>2</sup> < .001 | F(1,44) = 0.01, p =.921, partial eta<sup>2</sup> < .001 |

Notes. SEP: socioeconomic position. Foods with portion manipulation for Study 1: lunch (initial portion), dinner (initial portion), for Study 2: breakfast (initial portions), lunch (initial portion), dinner (initial portion). Non-manipulated foods: all other study food and self-reported food consumed.<sup>a</sup> Area under the curve of meal ratings taken before and after each meal, across entire day. SEP: socioeconomic position.

### 3.3 Effect of portion size on energy intake from foods with portion manipulation

In Study 1, there was a main effect of portion size condition on energy intake from portion-manipulated foods, with 249kcal less eaten from smaller (vs larger) lunch and dinner portion-manipulated servings (95% CI: 206, 292). There was no main effect or interaction with SEP (ps> .151). The same pattern of results was observed in Study 2; there was a main effect of portion size condition on energy intake from portion-manipulated foods, with 202kcal less eaten from smaller (vs larger) breakfast, lunch and dinner portion-manipulated servings (95% CI: 144, 259). There was no main effect or interaction with SEP (ps>.222).

### 3.4 Effect of portion size on energy intake from non-manipulated foods

In Study 1, there was no main effect of portion size condition on energy intake from non-manipulated foods (p = .772), a main effect of SEP with higher SEP consuming 414kcal more from non-manipulated foods than lower SEP (95%CI: 139, 688) (p = .004), and no interaction between portion size and SEP (p = .228). In Study 2, there was no main effect of portion size condition on energy intake from non-manipulated foods (p = .228), and no main effect or interaction with SEP (ps > .228).

### 3.5 Additional analyses

In Study 1, there was no evidence that portion size reduction impacted hunger or fullness ratings (planned), nor any evidence of SEP differences or SEP*portion size interactions; ps > .124. The same pattern of results was observed in Study 2, ps > .295. See Table 2 for descriptive statistics, and Table 3 for full ANOVA results. In both studies, foods tended to be well-liked and familiar to participants, with smaller and larger portions perceived to be ‘normal’ in size, and there was no evidence that rated liking, familiarity, or perceived normality of portions differed by SEP group in Study 1 or 2 (ps > .057). Less than half of participants reported noticing the portion manipulation and could accurately distinguish the portions they had on each day (Study 1: 44%; Study 2: 37%), with no evidence that this differed by SEP group. As we found no evidence of moderation of the effect of portion size on energy intake by SEP, we examined moderation by the measured individual difference measures (e.g., health and weight control food choice motives, satiety responsiveness, BMI) and found no evidence of moderation for any of the measures. See Supplementary Materials for secondary analyses in full.

## 4. Discussion

In two experiments we examined the impact of reducing food portion size on energy intake and found that when served smaller portions participants consumed less energy across the course of a day. Importantly, there was no evidence of moderation by SEP. The effect portion size had on energy intake was similar in participants from higher vs. lower SEP and findings were consistent across a range of SEP measurements, including education level, household income, childhood and self-perceived (subjective SEP).

Our findings are consistent with a large body of evidence showing that smaller portions decrease energy intake (13, 14) and this is consistent with recent research which suggests that changes to portion size are not fully compensated for over the course of a day (15, 31). The present results are not – however – consistent with findings from a recent study which found that lower SEP individuals were more susceptible to the portion size effect, intending to eat more from large portions of unhealthy snacks than higher SEP individuals in a hypothetical task (17). One limitation of this previous study is the use of hypothetical or ‘intended’ consumption and this may explain why our results differ. An additional explanation may be that the previous study findings of SEP moderation are limited only to discretionary snack foods and not meal energy intake, as examined in the present studies. In line with this, Best and Papies only found evidence of socioeconomic differences in susceptibility to the portion size effect for unhealthy (e.g., fries, cookies) but not healthy (e.g., carrot sticks, grapes) snacks (17), but did not examine meal energy intake. Future research could explore SEP differences in susceptibility to the portion size effect using a range of meals and snack foods to determine whether findings differ depending on food type.

In the present studies there was no evidence that energy intake from non-manipulated study foods (e.g., consuming more later in the day after receiving larger vs. smaller portions) differed by SEP. This suggests that individuals with lower SEP are no more susceptible to eating beyond energy needs after being served large meals than those with higher SEP, contrary to recent suggestions (18). In their study, Wijayatunga and colleagues provided participants with a very large lunch (60% of daily energy requirements) to be consumed in full and measured post-lunch energy intake. Conversely, the present studies provided smaller and larger portions deemed as ‘normal’ in size, and participants consumed as much as they liked *ad* libitum, which we presume is more representative of real-world eating occasions. It is worth noting, however, that (18) was a pilot study with a small sample and the authors note that replication with larger samples would be required to verify findings. In addition, in [18] there was no effect of SEP on daily energy intake, which makes it unclear to what extent SEP was associated with a meaningful impact (i.e., sustained) on energy intake in the study.

It has been suggested that reducing portion sizes of commercially available food could be an effective strategy to promote population health (13, 24). Our findings provide further support for this. Given concerns that dietary interventions do not exacerbate health inequalities, these findings support previous evidence that interventions which target the food environment (as opposed to the individual) are less likely to widen health inequalities as they do not rely on factors such as how motivated an individual is around their health to be effective (e.g., (25, 26)). In line with this, we collected a range of individual difference measures, including food choice motivation by health, ability to inhibit impulsive responses (e.g., for tempting food), and responsiveness to satiety signals. We reasoned these factors could moderate the influence of portion size on energy intake given evidence relating to satiety responsiveness in children (42, 43), and for impulsivity and perception of appropriate portion sizes in adults (17)). However, we found no evidence that variation in any of these measures moderated the influence of reducing portion size on energy intake, which is consistent with the proposal that portion size may be a universal driver of human energy intake (44).

There are several strengths and limitations to note. Findings were replicated across two studies, regardless of how SEP was measured, and were robust to a range of sensitivity analyses. We recruited a diverse sample in terms of SEP and this provided a strong test of the hypothesis that SEP moderates the influence of portion size on energy intake. The sample was predominantly white and although this is consistent with population demographics in the UK, our findings may not generalise to other ethnic groups. Importantly we only recruited females across both studies which may limit generalisability. However, given evidence that sex does not moderate the portion size effect (44) and studies providing evidence for potential SEP moderation of the influence of portion size sampled all (17) or mostly females (18), we presume that this exclusion criterion would not affect results. The controlled laboratory conditions used enabled precise measurement of energy intake, but efforts to replicate these findings in free-living conditions and in real-world food environments are now warranted, as the influence of portion size may be larger in real-life vs laboratory settings, see (45). Energy intake was measured for a day. Although the influence portion size has on daily energy intake has been shown to be similar over time (i.e., effects when measured during a single day are similar to longer durations) (15), confirming findings over longer duration would be valuable. A small proportion of participants consumed non-study foods as in (42), but analyses accounted for this as total daily energy intake included non-study foods. The present studies were powered to detect small-to-medium effects and although the interaction between SEP and portion size was not significant, if there is in fact a very small effect of SEP then we were not powered to detect this and very large sample sizes would be required to do so.

## Conclusions

Smaller meal portions reduced daily energy intake in both higher and lower SEP participants similarly. Reducing meal portion sizes could be an effective way to reduce overall daily energy intake and contrary to other suggestions it may be a socioeconomically equitable approach to improving diet

## Data Availability

All data produced are available online at

https://osf.io/scjn6/

https://osf.io/hx75k/

## Abbreviations

BMI: body mass index
PSE: portion size effect
SEP: socioeconomic position

## Acknowledgements

The authors would like to thank Alexandra Cunningham and Ahmed A. Sadab for their assistance with data collection.

## Statement of contributions

The authors’ responsibilities were as follows: For Study 1, TL, LM, AJ and ER designed the research, and for Study 2, all authors designed the research. For both studies, TL and KC conducted the research, and TL and AJ analysed the data. TL and ER wrote the paper and had primary responsibility for final content and all authors have read and approved the final manuscript.

## Data sharing

Data described in the manuscript will be made publicly and freely available without restriction at https://osf.io/scjn6/ and https://osf.io/hx75k/.

